# IL-1 Mediates Tissue Specific Inflammation and Severe Respiratory Failure In Covid-19: Clinical And Experimental Evidence

**DOI:** 10.1101/2021.04.09.21255190

**Authors:** Georgios Renieris, Eleni Karakike, Theologia Gkavogianni, Dionysia- Eirini Droggiti, Dionysios Kafousopoulos, Mihai G. Netea, Jesper Eugen-Olsen, John Simard, Evangelos J. Giamarellos-Bourboulis

## Abstract

**Background:** Acute respiratory distress syndrome (ARDS) in COVID-19 has been associated with dysregulated immune responses leading to catastrophic inflammation. The activation pathways remain to be fully elucidated. We investigated the ability of circulating to induce dysregulated immune responses.

**Materials & Methods:** Calprotectin and high mobility group box 1 (HMGB1) were associated with ARDS in 60 COVID-19 patients. In a second cohort of 40 COVID-19 patients calprotectin at hospital admission was associated with serum levels of soluble urokinase plasminogen activator receptor (suPAR). A COVID-19 animal model was developed by intravenous injection of plasma from healthy volunteers or patients with COVID-19 ARDS into C57/BL6 mice once daily for 3 consecutive days. In separate experiments, mice were treated with a) the IL-1 receptor antagonist Anakinra or vehicle and b) Flo1-2a anti-murine anti-IL-1α monoclonal antibody or the specific anti-human IL-1α antibody XB2001, or isotype controls. Mice were sacrificed on day 4. Cytokines and myeloperoxidase (MPO) in tissues were measured.

**Results:** Calprotectin, but not HMGB1, was elevated ARDS. Higher suPAR readouts indicated higher calprotectin levels. CHallenge of mice with COVID-19 plasma led to inflammatory reactions in murine lung and intestines as evidenced by increased levels of TNFα, IL-6, IFNγ and MPO. Anakinra treatment brought these levels down. Similar decrease was found in mice treated with Flo1-2a but not with XB2001.

**Conclusion:** Circulating alarmins, specifically calprotectin, of critically ill COVID-19 patients induces tissue-specific inflammatory responses through an IL-1α mediated mechanism. This could be attenuated through inhibition of IL-1 receptor or specific inhibition of IL-1α.

## INTRODUCTION

A little over one year since the onset of the COVID-19 pandemic, more than 2.3 million deaths have been attributed to the virus (https://www.who.int/emergencies/diseases/novel-coronavirus-2019). Despite all efforts to understand the disease, efforts are ongoing to elucidate what drives progression in COVID-19 cases from lower respiratory tract infections (LRTI) to acute respiratory distress syndrome (ARDS). We have suggested that the deterioration from pneumonia to ARDS is the result of a complex immune dysregulation, which in about 25% of patients appears to be related to a macrophage activation like syndrome; while in the remaining 75% of patients the mechanism seems to involve an interleukin (IL)-6 receptor dependent dysregulation, with an associated lymphopenia and a subset of cytokine over-producing monocytes that show decreased human leukocyte antigen (HLA)-DR expression (1).

Recent data suggests that SARS-CoV-2 stimulates vicious pro-inflammatory responses inducing NLRP3 inflammasome activation in monocytes and macrophages (2). In the open-label phase II study SAVE (3), patients with LRTI caused by SARS-CoV-2 and showing plasma concentrations of the biomarker suPAR (soluble urokinase plasminogen activator receptor) ≥ 6 ng/ml were treated with the recombinant human IL-1 receptor antagonist Anakinra. Anakinra is able to block both IL-1α and IL-1β bioactivity at the level of their common receptor (4) and prevent the development of ARDS.

The results of the SAVE trial led us to hypothesise that LRTI caused by SARS-CoV-2 can progress to ARDS. In this hypothesis, as a result of infection by SARS-CoV-2, patients over-produce IL-1α and other danger-associated molecular patterns (DAMPs) from their lung parenchyma. DAMPs trigger the release of IL-1β and in combination with IL-1α evoke strong innate immune pro-inflammatory responses which lead to ARDS (1). In order to test our hypothesis, we used a three-step approach. In the first step, we measured and found higher concentrations of the DAMP calprotectin in the plasma of patients with ARDS than patients without ARDS. In the second step, we measured and found that DAMP concentrations in the plasma of patients with suPAR more than 6 ng/ml were significantly higher than that in patients with suPAR less than 6 ng/ml. In the third step, we studied a COVID-19-like animal model, where we prevented the development of strong pro-inflammatory responses through inhibition of IL-1 signalling via Anakinra or anti-IL-1α antibodies.

## PATIENTS & METHODS

### Clinical study

Plasma samples from a cohort of 60 patients with and without ARDS that was determined to be caused by SARS-CoV (approval 30/20 by the National Ethics Committee of Greece; approval IS 021-20 by the National Organization for Medicines of Greece) and from 40 participants in the SAVE trial (suPAR-guided Anakinra treatment for Validation of the risk and Early management of severe respiratory failure by COVID-19; EudraCT number 2020-001466-11; National Ethics Committee approval 38/20; National Organization for Medicines approval ISO 28/20; ClinicalTrials.gov registration NCT04357366) were analyzed.

Patients were enrolled after written informed consent was provided by themselves or by first-degree relatives in case of patients unable to consent. All patients were adults of either sex with positive molecular testing of SARS-CoV-2 from respiratory secretions and with LRTI defined by the presence of diffuse infiltrates by chest X-ray or computed tomography of the chest. Exclusion criteria were a) HIV-1 infection and b) neutropenia defined as less than 1,000 neutrophils/mm^3^. Blood was sampled on the day of hospital admission in the emergencies. For patients admitted with ARDS, blood was collected within 24 hours from mechanical ventilation (MV). Control blood samples were collected from healthy volunteers matched for age and gender.

The following clinical variables were recorded: i) demographics; ii) severity scores, namely Acute Physiology and Chronic Health Evaluation (APACHE) II score, Charlson’s Comorbidity Index (CCI) and Sequential Organ Failure Assessment (SOFA) score; (iii) absolute blood cell counts, biochemistry, and blood gases; (iv) and progression into ARDS during the entire hospital stay. ARDS was defined as a ratio of partial oxygen pressure to fraction of inspired oxygen (PaO_2_ / FiO_2_) below 150 mmHg necessitating mechanical ventilation.

Five millilitres of whole blood was collected into a tube containing ethylenediaminetetraacetic acid (EDTA) and centrifuged. Calprotectin (S100A8/A9); high-mobility group box 1 (HMGB1); suPAR and ferritin were measured by enzyme-linked immunosorbent assays (ELISA) (InflaRx, Jena, Germany and Creative Diagnostics, Shirley, NY, USA; suPARnostic, ViroGates, Lyngby, Denmark and ORGENTEC Diagnostika GmbH, Mainz, Germany, respectively); and C-reactive protein (CRP) was measured using a nephelometric assay (Behring, Berlin, Germany). The lowest limits of detection were: 0.5 ng/ml for HMGB1; 1.1 ng/ml for suPAR; 0.2 mg/l for CRP; and 75 ng/ml for ferritin. RNA isolation of SARS-CoV-2 was performed with the QIAamp Viral RNA Mini Kit (Qiagen, Hilden, Germany). Reverse transcription of the viral RNA into cDNA and enrichment of the sequences of interest via Real-time Polymerase Chain Reaction (PCR) was performed in the Rotor-Gene Q instrument (Qiagen) using the SARS-CoV-2 reagent Viasure (CerTest Biotec SL, Zaragoza, Spain).

### Animal model of COVID-19 Pathogenic Inflammation

Animal experiments were conducted in the Unit of Animals for Medical and Scientific purposes of the University General Hospital “Attikon” (Athens, Greece). All experiments were licensed from the Greek veterinary directorate under the protocol number 471955/06-07-2020.

We studied 80 male and female C57Bl6 mice (7-8 weeks old). Mice were allowed to acclimate for seven days before beginning of the experiments. They were housed in typical mouse cages, up to 5 mice per cage on 12-h dark/light cycle and allowed free access to standard dry rodent diet and water. Analgesia was achieved with paracetamol suppositories to avoid interactions with the immune system.

In order to develop a model of COVID-19-like inflammatory illness, mice were treated intravenously (iv) with 100 μl of plasma from patients with ARDS due to SARS-CoV-2 infection for 3 consecutive days; control mice were treated with 100 μl of plasma from healthy volunteers for 3 consecutive days. On the fourth day, mice were sacrificed by subcutaneous (sc) injection of 300 mg/kg ketamine, followed by cervical dislocation. Under sterile conditions a midline abdominal incision was performed. Then, segments of liver, lower lobe of the right lung, right kidney, ileum (ca. 1 cm distally to the pyloric sphincter) and colon (ca. 1 cm proximally of the anal ring) were excised and collected into sterile tubes with 1 ml NaCl 0.9%. The intestinal samples were thoroughly flushed with NaCl 0.9% for removal of feces before storage. The samples were weighed and homogenized.

Segments of spleens were gently squeezed and passed through a sterile filter (250 mm, 12-13 cm, AlterChem Co, Athens, Greece) for the collection of splenocytes. Peritoneal cells were retrieved by peritoneal lavage with 10 ml of cold PBS containing 2 mmol/l of EDTA. After three serial washings, spleencells and peritoneal cells were counted on Neubauer plates with trypan blue for exclusion of dead cells. A total of 5 × 10^6^ cells/ml were incubated in sterile 24-well plates in RPMI-1640, supplemented with 2 mM glutamine, 10% fetal bovine serum, 100 U/ml of penicillin G and 0.1 mg/ml of streptomycin, in the absence or presence of 10 ng/ml lipopolysaccharide (LPS) of *Escherichia coli* O55:B5 or 5 × 10^5^ cfu/ml heat killed *Candida* α*lbicans (*HKCA). After 24 hours or 5 days of incubation at 37°C in 5% CO_2_, the plates were centrifuged, and the supernatants were collected. Concentrations of tumor necrosis factor alpha (TNFα), interferon gamma (IFNγ) and IL-6, IL-17A and IL-22 in supernatants from tissue samples and spleencells were measured in duplicate by an enzyme immunosorbent assay (ThermoFisher Scientific, Massachussets, USA) according to the manufacturer’s instructions. The lowest detections limits were as follows: for TNFα 19 pg/ml; for IFNγ 16 pg/ml; for IL-6 10 pg/ml; for IL-17A 8 pg/ml; and for IL-22 16 pg/ml.

Myeloperoxidase (MPO) activity in all collected tissues was determined. Tissue segments were homogenized with T-PER® (ThermoFisher Scientific) and centrifuged at 10,000 rpm at 4°C. Then the homogenates were incubated in wells of a 96-well plate at 37°C with 4.2 mM tetramethylbenzidine (Serva, Heidelberg, Germany), 2.5 mM citrate, 5 mM NaH_2_PO_4_ and 1.18 mM H_2_O_2_ pH 5.0 at a final volume of 150 μl. After 5 minutes the reaction was terminated by adding 50 ml 0.18 M H_2_SO_4_. Absorbance was read at 450 nm against blank wells. Results were adjusted for tissue sample protein content on Bradford assay (Sigma-Aldrich) and they were expressed as MPO units/mg protein/g.

Experiments were repeated in mice receiving in addition to i.v. plasma, a) 100 μl sc of 10 mg/kg Anakinra (Swedish Orphan Biovitrum, Stockholm, Sweden) or 100 μl of 0.9% NaCl once daily for 3 days; and b) 100 μl intraperitoneally (ip) of 1000 μg/ml (12) anti-murine IL-1α antibody Flo1-2a or human IL-1α antibody XB2001, or murine isotype control (XBiotech, TX, USA).

In a separate experiment, mice treated with COVID-19 plasma for three days were challenged on the fourth day ip with 1⨯10^7^ cfu/ml *E. coli* or *Acinetobacter baumannii*. Survival was recorded every 12 hours for 7 days.

### Statistics

Categorical data were presented as frequencies and quantitative variables as mean ± SE. Comparisons between groups were done using the Fisher exact test for categorical data. Comparison for quantitative data was performed using the Mann-Whitney U test for two group comparisons and the one-way ANOVA with the Bonferroni correction for multiple group comparison. Correlations were performed using the Spearman’s rank of order. Any p value below 0.05 was considered statistically significant.

## RESULTS

Forty consecutive patients with COVID-19 who did not develop ARDS during their hospital stay and 20 consecutive patients with ARDS were analyzed. Their demographics are shown in Table 1. Samples of 10 healthy volunteers were analyzed as healthy controls. Serum calprotectin levels were significantly higher among patients with COVID-19 and ARDS (Figure 1A). HMGB1 concentrations did not differ between patients with COVID-19 with and without ARDS (Figure 1B). Elevated calprotectin levels COVID-19-related ARDS raised the question whether calprotectin is released early among patients and may serve as a marker for patients who will progress to ARDS.

**Table 1.** Baseline clinical and laboratory characteristics of patients according to the development of ARDS due to pneumonia by the SARS-CoV-2 coronavirus.

|  | No ARDS (n=40) | ARDS (n=20) | p-value |
| --- | --- | --- | --- |
| Age (years, mean ± SD) <sup>#</sup> | 57.3 ± 14.85 | 66.60 ± 11.45 | 0.020<sup>#</sup> |
| Male gender (n, %) | 33 (82.5) | 17 (85.0) | 0.559<sup>□</sup> |
| CCI (mean ± SD) <sup>#</sup> | 1.85 ± 1.66 | 3.55 ± 2.56 | 0.012<sup>#</sup> |
| APACHE II score (mean ± SD) <sup>#</sup> | 5.50 ± 3.11 | 10.05 ± 5.07 | 0.001<sup>#</sup> |
| SOFA (mean ± SD) <sup>#</sup> | 1.87 ± 1.66 | 3.35 ± 1.06 | 0.001<sup>#</sup> |
| Main comorbidities (n, %) |  |  |  |
| Type 2 diabetes mellitus | 6 (15.0) | 6 (30.0) | 0.189<sup>□</sup> |
| Congestive heart failure | 1 (2.5) | 2 (10.0) | 0.255<sup>□</sup> |
| Coronary heart disease | 1 (2.5) | 4 (20.0) | 0.038<sup>□</sup> |
| Chronic renal disease | 1 (2.5) | 0 (0) | 0.667<sup>□</sup> |
| COPD | 4 (10.0) | 3 (15.0) | 0.676<sup>□</sup> |
| Laboratory values (mean ± SD) | | | |
| White blood cells (mm<sup>3</sup>) | 6098.46 ± 2102.44 | 8758.00 ± 3992.08 | 0.003<sup>#</sup> |
| Absolute neutrophil counts (mm<sup>3</sup>) | 4535.52 ± 2110.35 | 7437.75 ± 3762.13 | 0.001<sup>#</sup> |
| Absolute lymphocyte counts (mm<sup>3</sup>) | 1065.98 ± 419.86 | 810.04 ± 426.46 | 0.025<sup>#</sup> |
| Platelets (x 10<sup>3</sup>, mm<sup>3</sup>) | 191.2 ± 85.6 | 178.3 ± 54.9 | 0.389<sup>#</sup> |
| C-reactive protein (mg/l) | 124.55 ± 100.73 | 206.38 ± 85.96 | 0.014<sup>#</sup> |
| Fibrinogen (mg/dl) | 510.59 ± 172.91 | 611.20 ± 199.78 | 0.262<sup>#</sup> |
| Ferritin (ng/ml) | 1136.05 ± 1022.05 | 1288.83 ± 573.13 | 0.381<sup>#</sup> |
| Procalcitonin (ng/ml) | 0.25 ± 0.39 | 0.38 ± 0.46 | 0.280<sup>#</sup> |
| D-dimers (ng/ml) | 599.76 ± 364.72 | 763.00 ± 702.42 | 0.464<sup>#</sup> |
| AST (U/l) | 41.32 ± 26.62 | 64.80 ± 108.08 | 0.502<sup>#</sup> |
| ALT (U/l) | 39.33 ± 34.24 | 52.80 ± 74.30 | 0.822<sup>#</sup> |
| Creatinine (mg/dl) | 0.89 ± 0.18 | 1.29 ± 1.16 | 0.006<sup>#</sup> |
| Total bilirubin (mg/dl) | 0.59 ± 0.54 | 5.25 ± 18.34 | 0.211<sup>#</sup> |

Abbreviations: SD: standard deviation; APACHE: acute physiology and chronic health evaluation; CCI: Charlson's comorbidity index; SOFA: sequential organ failure; COPD: chronic obstructive pulmonary disease; AST: aspartate aminotransferase; ALT: alanine aminotransferase; ARDS: acute respiratory distress syndrome
<sup>□</sup>Comparison by Fischer exact test; <sup>#</sup>Comparison by the Mann-Whitney U test.

**Figure 1.**
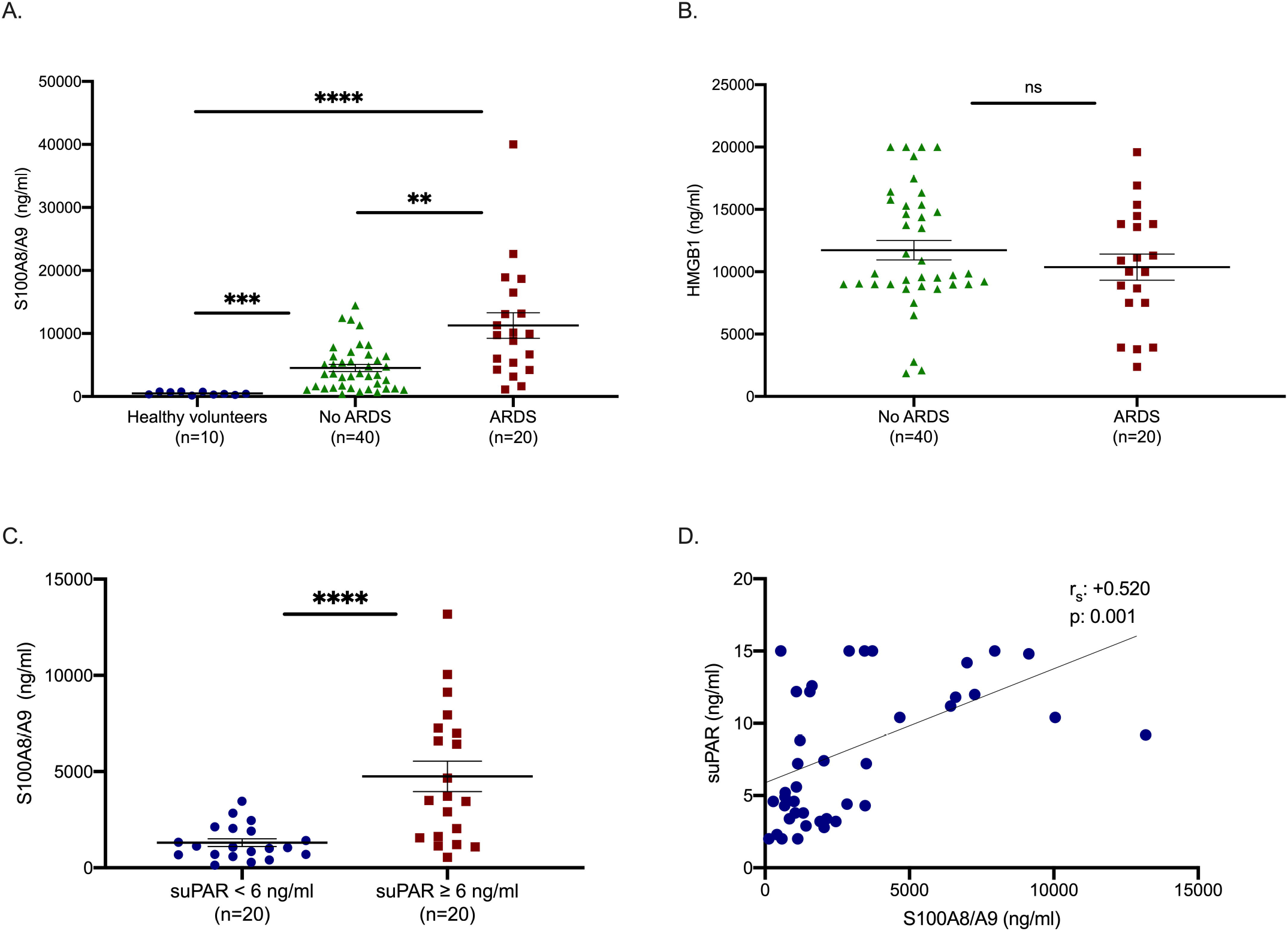
Calprotectin is increased in patients with ARDS due to COVID-19. A-B) Concentrations of calprotectin (S100A8/A9) and high mobility group box 1 (HMGB1) were measured in the plasma from healthy volunteers; from patients with COVID-19 who did not develop ARDS during follow-up on the day of hospital admission; and from patients with ARDS by COVID-19 on the first day of ARDS. Comparison by the Mann Whitney U test; ns non-significant; ^⍰^ p< 0.05; ^⍰⍰^ p< 0.01; ^⍰⍰⍰^ p< 0.001; ^⍰⍰⍰⍰^; p< 0.0001. Panels C and D) suPAR was measured in 40 patients screened for participation in the SAVE trial (3). None of the patients with suPAR less than 6 ng/ml developed acute respiratory distress syndrome (ARDS), whereas 6 patients (30%) with suPAR 6 ng/ml or more developed ARDS. C) Concentrations of calprotectin in 20 patients with suPAR less than 6 ng/ml and in 20 patients with suPAR 6 ng/ml or more. Comparison by the Mann Whitney U test; ^⍰^ p< 0.05. D) Correlation of levels of suPAR with plasma calprotectin on day of admission Spearman rank correlation coefficients (r_s_), interpolation lines and p-values are provided.

suPAR was previously identified as an early predictor of ARDS progression in patients with at least 6 ng/ml serum levels in the SAVE trial (3). To study of the early increase of suPAR denotes increases of calcprotectin, calprotectin was measured in 40 patients screened for inclusion in SAVE trial. 20 patients were with suPAR concentrations less than 6 ng/ml and 20 patients showed suPAR concentrations ≥ 6 ng/ml (Table 2). None of the patients with suPAR less than 6 ng/ml developed ARDS; 30% of those with suPAR ≥ 6 ng/ml progressed into ARDS. Remarkably, patients with suPAR concentrations ≥ 6 ng/ml had significantly higher concentrations of calprotectin (Figure 1C). A significant positive correlation between calprotectin and suPAR was thus demonstrated (Figure 1D). These findings demonstrated that both suPAR and calprotectin are reliable prognostic biomarkers in predicting the development of severe respiratory failure in COVID-19. These findings also suggested that DAMPs like calprotectin, released at early stages of clinical infection are involved in mediating the progression of pneumonia into ARDS.

**Table 2.** Baseline clinical and laboratory characteristics of patients with pneumonia by SARS-CoV-2 coronavirus according to the measured levels of circulating soluble urokinase plasminogen activator receptor (suPAR)

|  | suPAR < 6 ng/ml<br>(n=20) | suPAR ≥ 6 ng/ml<br>(n=20) | p-value |
| --- | --- | --- | --- |
| Age (years, mean ± SD) <sup>#</sup> | 46.25 ± 17.63 | 68.30 ± 9.27 | 6 x 10<sup>-5#</sup> |
| Male gender (n, %) | 13 (65.0) | 13 (65.0) | 1.000<sup>□</sup> |
| CCI (mean ± SD) <sup>#</sup> | 3.56 ± 1.62 | 5.13 ± 0.99 | 0.022<sup>#</sup> |
| APACHE II score (mean ± SD) <sup>#</sup> | 7.28 ± 3.43 | 6.69 ± 5.25 | 0.953<sup>#</sup> |
| SOFA (mean ± SD) <sup>#</sup> | 0.86 ± 1.07 | 2.61 ± 1.20 | 0.004<sup>#</sup> |
| ARDS (n, %) | 0 (0.0) | 6 (30) | 3 x 10<sup>-6□</sup> |
| Main comorbidities (n, %) |  |  |  |
| Type 2 diabetes mellitus | 1 (5.0) | 4 (20.0) | 0.342<sup>□</sup> |
| Congestive heart failure | 0 (0) | 1 (5.0) | 0.500<sup>□</sup> |
| Coronary heart disease | 1 (5.0) | 3 (15.0) | 0.658<sup>□</sup> |
| COPD | 0 (0) | 1 (5.0) | 0.506<sup>□</sup> |
| Laboratory values (mean ± SD) |  |  |  |
| White blood cells (mm<sup>3</sup>) | 6007.14 ± 1500.55 | 7578.88 ± 3201.64 | 0.220<sup>#</sup> |
| Absolute neutrophil counts (mm<sup>3</sup>) | 3573.30 ± 1723.14 | 5833.80 ± 2988.62 | 0.055<sup>#</sup> |
| Absolute lymphocyte counts (mm<sup>3</sup>) | 2233.86 ± 1872.39 | 1203.26 ± 614.70 | 0.198<sup>#</sup> |
| Platelets (x 10<sup>3</sup>, mm<sup>3</sup>) | 261.8 ± 133.8 | 212.1 ± 68.2 | 0.574<sup>#</sup> |
| C-reactive protein (mg/l) | 35.69 ± 53.43 | 89.19 ± 82.45 | 0.083<sup>#</sup> |
| Fibrinogen (mg/dl) | 352.00 ± 59.24 | 612.22 ± 187.67 | 0.003<sup>#</sup> |
| Ferritin (ng/ml) | 283.41 ± 244.73 | 882.90 ± 595.75 | 0.024<sup>#</sup> |
| Procalcitonin (ng/ml) | 0.06 ± 0.05 | 0.36 ± 0.36 | 0.133<sup>#</sup> |
| D-dimers (ng/ml) | 421.67 ± 324.10 | 1407.79 ± 3251.55 | 0.207<sup>#</sup> |
| AST (U/l) | 22.57 ± 6.68 | 46.44 ± 29.49 | 0.041<sup>#</sup> |
| ALT (U/l) | 27.14 ± 14.37 | 38.67 ± 27.78 | 0.458<sup>#</sup> |
| Creatinine (mg/dl) | 0.92 ± 0.27 | 0.97 ± 0.34 | 0.883<sup>#</sup> |
| Total bilirubin (mg/dl) | 0.259 ± 0.11 | 1.54 ± 2.06 | 0.001<sup>#</sup> |

Abbreviations: SD: standard deviation; APACHE: acute physiology and chronic health evaluation; CCI: Charlson's comorbidity index; SOFA: sequential organ failure; ARDS: acute respiratory distress syndrome; COPD: chronic obstructive pulmonary disease; AST: aspartate aminotransferase; ALT: alanine aminotransferase.
<sup>□</sup>Comparison by Fischer exact test; <sup>#</sup>Comparison by the Mann-Whitney U test.

In order to investigate the mechanism by which calprotectin induces the hyperinflammation related to COVID-19 associated respiratory failure, we sought to design a COVID-19 murine model. Mice were challenged with plasma either from healthy volunteers (HV) or from patients with ARDS of COVID-19. Plasma of COVID-19 patients led to organ-specific increase of cytokine concentrations. More precisely, TNFα and MPO activity were specifically increased in the lung, ileum and colon of mice challenged with COVID-19 plasma; IL-6 was upregulated in the lung and colon; and IFNγ was upregulated in the lung (Figure 2A to D). These findings demonstrated a compartmentalized inflammatory response in mice after challenging with COVID-19 patient plasma.

**Figure 2.**
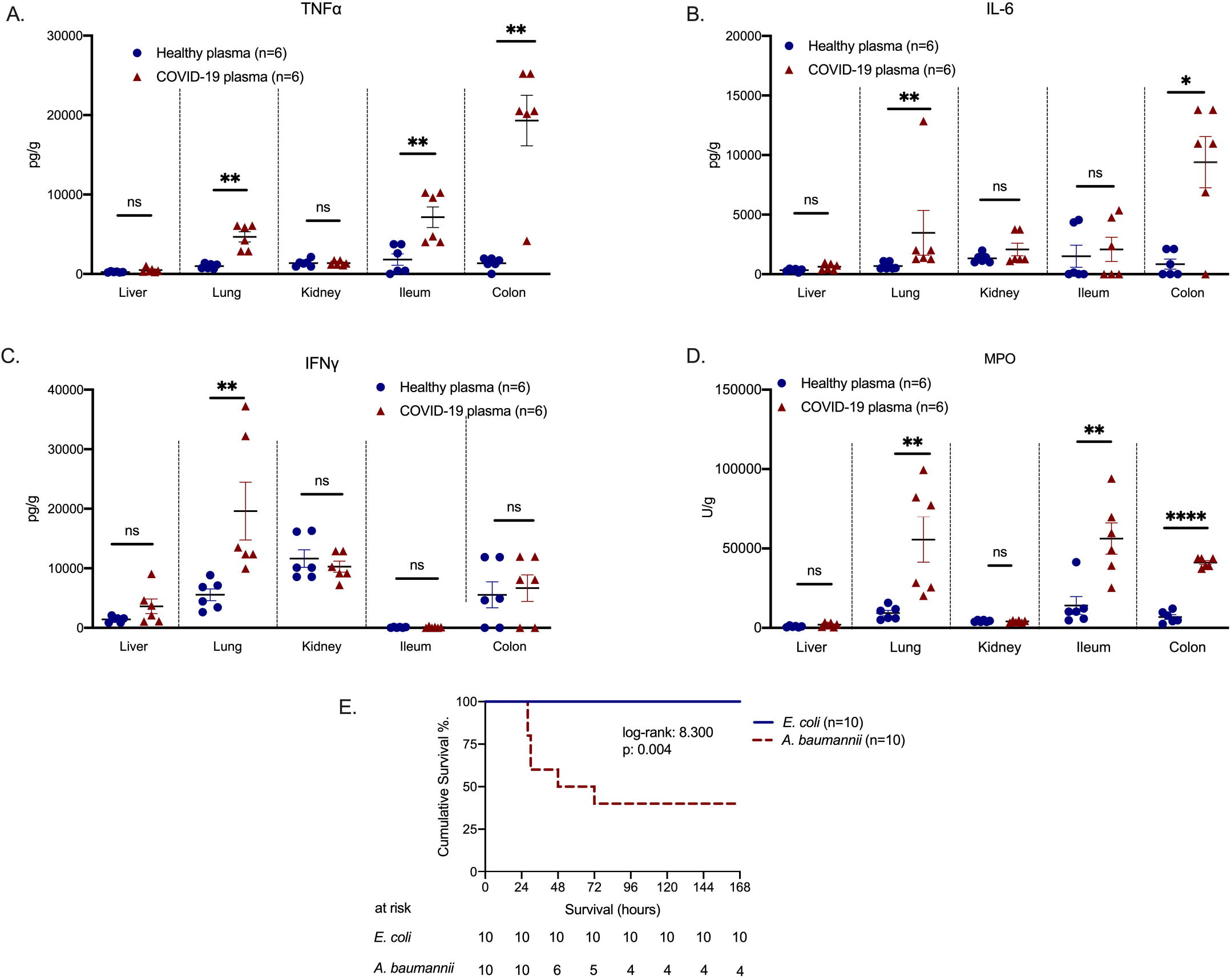
A COVID- like animal model of compartmentalized hyperinflammation. C57Bl6 mice were injected intravenously (i.v.) with plasma of healthy volunteers or patients with ARDS due to COVID-19 for three consecutive days. Mice were sacrificed on day 4. Tissue concentrations of A) tumor necrosis factor alpha (TNFα); B) Interleukin (IL)-6; C) Interferon gamma (IFNγ) and D) myeloperoxidase (MPO) activity were determined. Comparison by the Mann Whitney U test; ns non-significant; ^⍰^ p< 0.05; ^⍰⍰^ p< 0.01; ^⍰⍰⍰⍰^ p< 0.0001. E) C57Bl6 mice were injected intravenously (i.v.) with plasma of healthy volunteers or patients with ARDS due to COVID-19 for three consecutive days. On day 4 mice were challenged intraperitoneally (i.p.) with *E. coli* or *A. baumannii*. Survival comparison by the log-rank test and the respective p-value are provided.

DAMPs, like calprotectin, act on monocytes and macrophages and tend to stimulate intense IL-1 responses. To provide evidence on the contribution of IL-1 stimulation, mice challenged with plasma from HV or COVID-19 patients were treated with N/S or with Anakinra; the latter blocks both human and murine IL-1α and IL-1β. Treatment with Anakinra significantly reduced tissue concentrations of TNFα, IL-6, IFN-γ and MPO activity (Figure 3A to 3I). However, the observed inflammation may have been mediated through human IL-1α present in the plasma of COVID-19 patients. To further clarify this, mice challenged with COVID-19 plasma were treated with the anti-murine specific anti-IL-1α antibody Flo1-2a and with the human specific anti-human IL-1α antibody XB2001, respectively. Treatment with Flo1-2a significantly reduced tissue concentrations of TNFα, IL-6, IFNγ and MPO activity in contrast to treatment with XB2001 (Figure 4A to 4I). This suggests that the host-derived mouse IL-1α and not any injected human IL-1α was necessary to induce the compartmentalized inflammation.

**Figure 3.**
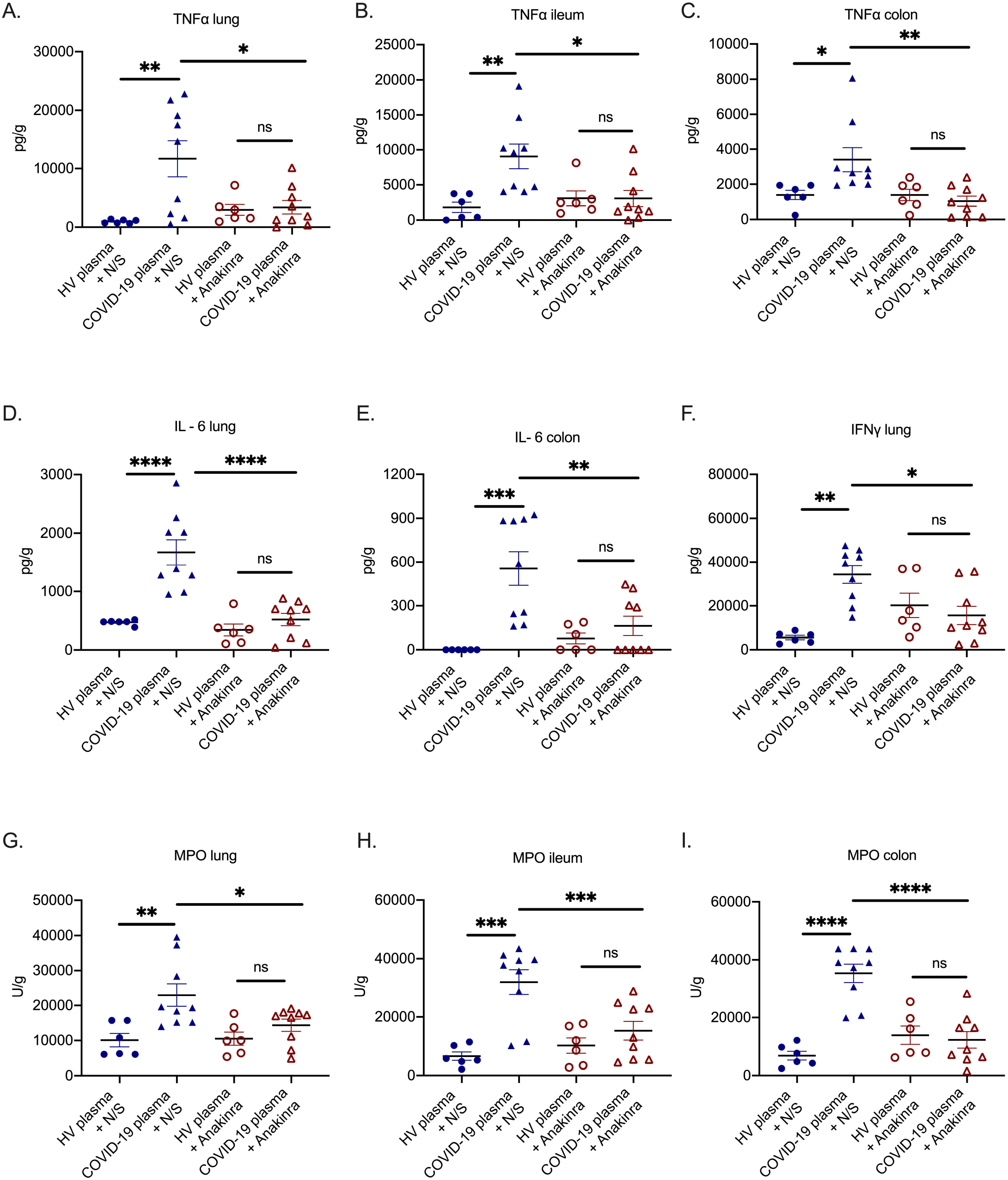
IL-1 inhibition attenuates the compartmentalized hyperinflammation in a COVID-like murine model. In a COVID-like infection model, C57Bl6 mice were challenged intravenously (i.v.) with plasma of healthy volunteers or patients with ARDS due to COVID-19 for three consecutive days. In separate experiments, on each day of plasma challenge, mice were treated with Anakinra, which inhibits human and murine IL-1, or vehicle. Mice were sacrificed on day 4. A-C) Tumor necrosis factor alpha (TNFα); D-E) Interleukin (IL) −6; F) Interferon gamma (IFNγ) and G-I) myeloperoxidase (MPO) activity was determined in tissues. Comparison by the Mann Whitney U test; ns non-significant; ^⍰^ p< 0.05; ^⍰⍰^ p< 0.01.

**Figure 4.**
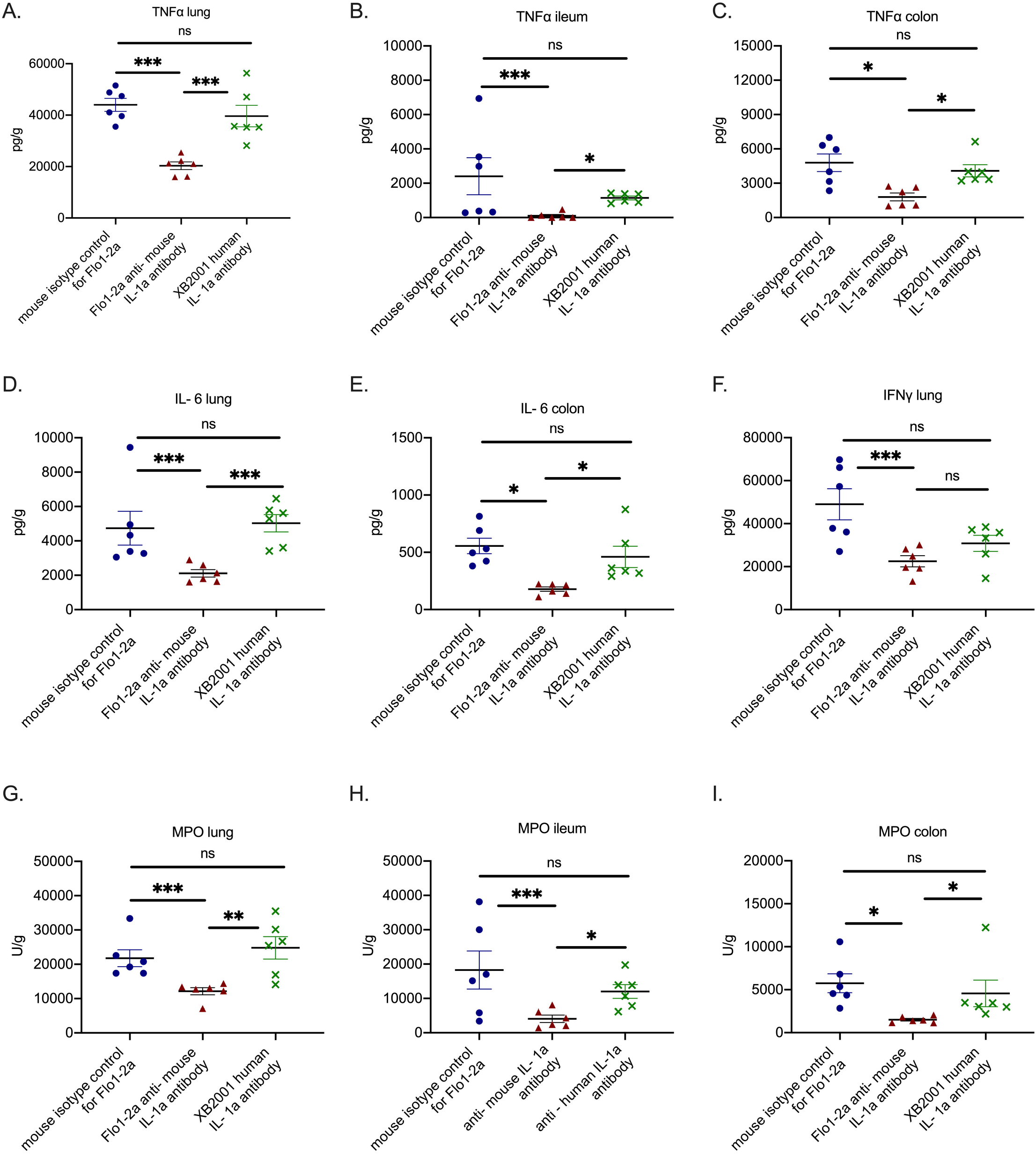
Murine IL-1α drives the hyperinflammation caused by SARS-CoV-2. In a COVID-like infection model, C57Bl6 mice were challenged intravenously (i.v.) with plasma of patients with ARDS due to COVID-19 for three consecutive days. In separate experiments, on each day of plasma challenge, mice were treated with Flo1-2a anti-murine IL-1α antibody or XB2001 human IL-1α antibody or murine isotype control for Flo1-2a. Mice were sacrificed on day 4. A-C) Tumor necrosis factor alpha (TNFα); D-E) Interleukin (IL) −6; F) Interferon gamma (IFNγ) and G-I) myeloperoxidase (MPO) activity was determined in tissues. Comparison by the Mann Whitney U test; ns non-significant; ^⍰^ p< 0.05; ^⍰⍰^ p< 0.01; ^⍰⍰⍰^ p< 0.001.

Treatment with Anakinra led to an increase in ex vivo production capacity of IFNγ and TNFα, but not of IL-6, IL-17A or IL-22 from splenocytes following stimulation with HKCA (Figure 5A to 5F and Figure S1A to S1D). HKCA targets lymphocyte responses from splenocytes and the cytokine responses are compatible with increase of Th1 responses following treatment with Anakinra.

**Figure 5.**
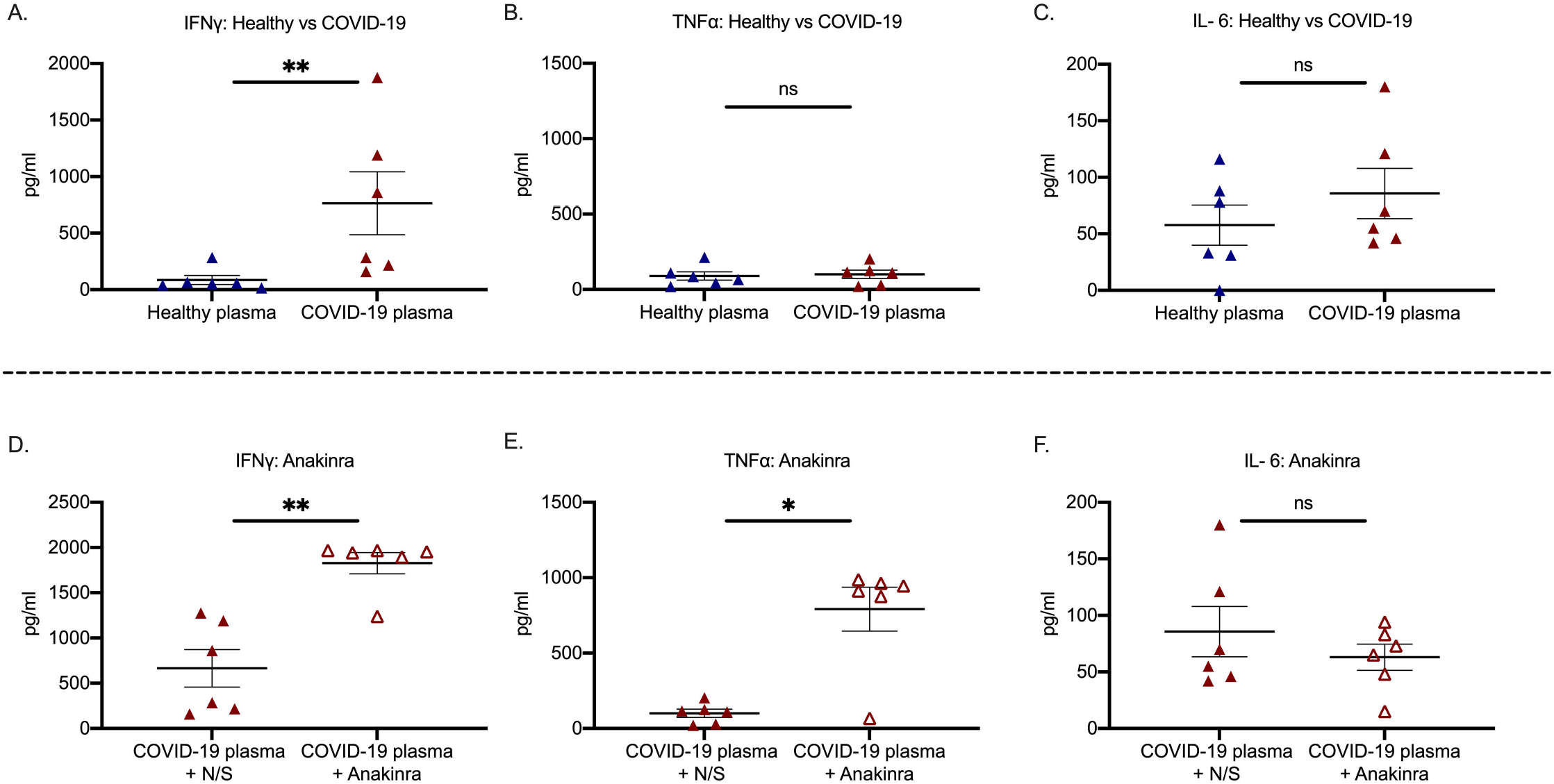
IL-1 inhibition primes Th1 responses in COVID-19. A-C) In a COVID-like infection model, C57Bl6 mice were treated intravenously (i.v.) with plasma of healthy volunteers or patients with ARDS due to COVID-19 for three consecutive days. Mice were sacrificed on day 4. Splenocytes were isolated and incubated with heat killed *C. albicans* for the production of interferon-gamma (IFNγ), tumour necrosis factor-alpha (TNFα) and interleukin (IL)-6. D-F) In a COVID-like infection model, C57Bl6 mice were treated intravenously (i.v.) with plasma of patients with ARDS due to COVID-19 with or without treatment with the IL-1 receptor inhibitor Anakinra for three consecutive days. Mice were sacrificed on day 4. Splenocytes were isolated and incubated with heat killed *C. albicans* for the production of IFNγ, TNFα and IL-6. Comparison by the Mann Whitney U test; ns non-significant; ^⍰^ p< 0.05; ^⍰⍰^ p< 0.01.

The number of transcripts of core SARS-CoV-2 genes *ORF1ab* and *N* were indirectly provided by the cycles (Ct) of PCR positivity in plasma samples that were used for the induction of the COVID-19-like hyperinflammation in the murine model. No correlation was found between Cts and the levels of TNFα, IL-6, IFNγ and MPO in the lung of COVID-19 patients with ARDS due to COVID-19 (Figure S2). This suggests that the hyperinflammation is independent from the viral load. In order to mimic hospital acquired secondary bacterial infections among COVID-19 patients in murine, mice treated for three consecutive days with COVID-19 plasma were further challenged with *A. baumannii or E. coli*. Survival was significantly lower after challenging with *A. baumannii* (Figure 3E). Observations coming from our prospective cohort of 134 ARDS patients revealed higher incidence of secondary infections by A. *baumannii* when compared to other Gram-negative bacteria (Table S1, Figure S3). This further supports the immunological similarities of the developed animal model to the human infection.

## DISCUSSION

This study presented evidence that severe respiratory failure in COVID-19 patients is possibly triggered by DAMPs, most likely calprotectin, which are released early after the interaction of SARS-CoV-2 with host cells. It may well be the case that calprotectin is this specific DAMP since circulating levels of HMGB1, another DAMP, do not follow those of calprotectin. DAMPs lead to a compartmentalized inflammation through enhancement of the production of pro-inflammatory cytokines IL-1α and IL-1β.

It has been clearly demonstrated that an impaired immune response is a core characteristic of patients who develop ARDS due to SARS-CoV-2 (5). The associated great increase in the mortality of these patients has turned the focus on potential therapeutic targets to disrupt the immune dysregulation. This necessitates a better understanding of the pathogenesis of COVID-19.

Calprotectin, a heterodimer of S100A8 and S100A9, is a protein secreted by leukocytes. The upregulation of calprotectin expression has been associated with various inflammatory processes such as inflammatory bowel disease, rheumatoid arthritis, spondyloarthritis and cardiovascular disease in type 2 diabetes mellitus (6-9). Additionally, calprotectin is crucial for neutrophil accumulation and inflammation in the lung parenchyma during bacterial community acquired pneumonia (10). Recently, high levels of calprotectin in samples of COVID-19 patients were correlated with disease severity and were associated with increased levels of inflammatory cytokines and chemokines and myelopoiesis of abnormal myeloid cells (11). In this study, we suggest calprotectin as a potential first element in the pathway leading to ARDS in COVID-19 patients. Indeed, we found greater levels in patients with ARDS compared to patients without ARDS. On the contrary, HMGB1, an additional alarmin, which has been associated with sepsis pathogenesis and outcome (12), did not significantly differ among patients with and without ARDS due to COVID-19. Moreover, suPAR which is an early predictor of respiratory deterioration in COVID-19 (3, 13) was indicative of patients with higher calprotectin levels, suggesting that increased suPAR can be the biomarker of DAMP-induced pathways in COVID-19.

SARS-CoV-2 uses the cellular surface protein angiotensin-converting enzyme 2 (ACE2) to bind and enter cells. However, the mouse ACE2 does not effectively bind the viral spike protein (14). Therefore wild-type mice cannot be directly used as a model to gain insight in COVID-19 pathophysiology. Mice over-expressing ACE2 have been used as a strategy to tackle this problem (15). While it is accepted that the interaction of the spike protein of SARS-CoV-2 with human angiotensin converting enzyme 2 (ACE2) is the first step for viral invasion in host cells of the upper and lower respiratory tract (16), the ability of the virus to directly infect other human tissues, where ACE2 is also expressed, such as the small intestine, kidneys, and the heart, is a matter of dispute (17). In addition, we also need to understand the pure immunological mechanisms of hyperinflammation in COVID-19. This is introducing the need of COVID-19 models like the one developed after animal challenge with human plasma. The model simulates the inflammatory reaction in human SARS-CoV-2 infection since it provides compartmentalized hyperinflammation of the lung.

Calprotectin acts through the Toll like receptor 4 (TLR4) and the receptor for advanced glycation end-products (RAGE) and may prime for intracellular pro-IL-1β (18) which is subsequently cleaved by SARS-CoV-2 activated NLRP3 inflammasome (2) into IL-1β. Anakinra prevents the inflammatory action of both IL-1α and IL-1β by blocking their cellular receptor. IL-1α released during cell death is characterized by an autocrine function enabling an amplifying loop of inflammation (19). Moreover, we have shown that Anakinra enhances the Th1 response in mice and probably improves viral clearance.

Our findings provide evidence that progression from LRTI by SARS-CoV-2 to ARDS is a process which is mediated by DAMPs, like IL-1α and calprotectin, released following cell destruction. The early detection of the presence of circulating DAMPs and blockade through IL-1a inhibition may represent a viable therapeutic strategy. This is the hallmark of the efficacy of Anakinra - guided by the biomarker suPAR - which led to a significant decrease in the incidence of ARDS in the SAVE trial (3).

## Data Availability

Data of this study are available after communication with the corresponding author.

## Acknowledgments

We thank Zhongli Xu, Katja Schubert, Marlen Hermann (InflaRx GmbH, Jena, Germany) for the measurements of calprotectin in patient samples as well as for their critical review of the manuscript.

## Competing interests

J. Eugen-Olsen is a cofounder, shareholder and CSO of ViroGates A7S, Denmark and is named inventor on patents on suPAR owned by Copenhagen University Hospital Hvidovre, Denmark. He is granted by the Horizon 2020 European Grant RISKinCOVID.

J. Simard is the CEO and founder of XBiotech.

M.G. Netea is a scientific founder of TTxD, and received research grants from GSK and ViiV Healthcare.

E.J. Giamarellos-Bourboulis has received honoraria from AbbVie USA, Abbott CH, InflaRx GmbH, MSD Greece, XBiotech Inc. and Angelini Italy; independent educational grants from AbbVie, Abbott, Astellas Pharma Europe, AxisShield, bioMérieux Inc, InflaRx GmbH, and XBiotech Inc.; and funding from the FrameWork 7 program HemoSpec (granted to the National and Kapodistrian University of Athens), the Horizon2020 Marie-Curie Project European Sepsis Academy (granted to the National and Kapodistrian University of Athens), and the Horizon 2020 European Grant ImmunoSep (granted to the Hellenic Institute for the Study of Sepsis).

## Funding

The study was funded in part by the Hellenic Institute for the Study of Sepsis and in part by the Horizon 2020 grant RISKinCOVID. The funders had no role in the design, collection, analysis and interpretation of data, writing, and decision to publish.

## Notes

### Clinical Trial

NCT04357366

### Author Declarations

National Ethics Committee of Greece and National Organization for Medicines of Greece

### Summary of Updates

3 of the original co-authors Zhongli Xu, Katja Schubert, Marlen Hermann have been removed from the author list and are acknowledged in the respective section off the manuscript

