## Supplementary figures and table for "IL-1 Mediates Tissue Specific Inflammation and Severe Respiratory Failure In Covid-19: Clinical And Experimental Evidence"

**Running heading: IL-1 bioactivity in COVID-19**

**Georgios Renieris^1^, Eleni Karakike^1^, Theologia Gkavogianni^1^, Dionysia- Eirini Droggiti^1^, Dionysios Kafousopoulos^1^, Mihai G. Netea^2,3^, Jesper Eugen-Olsen^4^, John Simard^5^, Evangelos J. Giamarellos-Bourboulis^1^**

**^1^ 4^th^ Department of Internal Medicine, National and Kapodistrian University of Athens, Medical School, 124 62 Athens, Greece**

**^2^Immunology and Metabolism, Life & Medical Sciences Institute, University of Bonn, 53115 Bonn, Germany**

**^3^Department of Internal Medicine and Center for Infectious Diseases, Radboud University, 6500 Nijmegen, the Netherlands**

**^4^Department of Clinical Research, Copenhagen University Hospital Hvidovre, 2650 Hvidovre, Denmark**

**^5^XBiotech, 78744 Austin, Texas, USA**

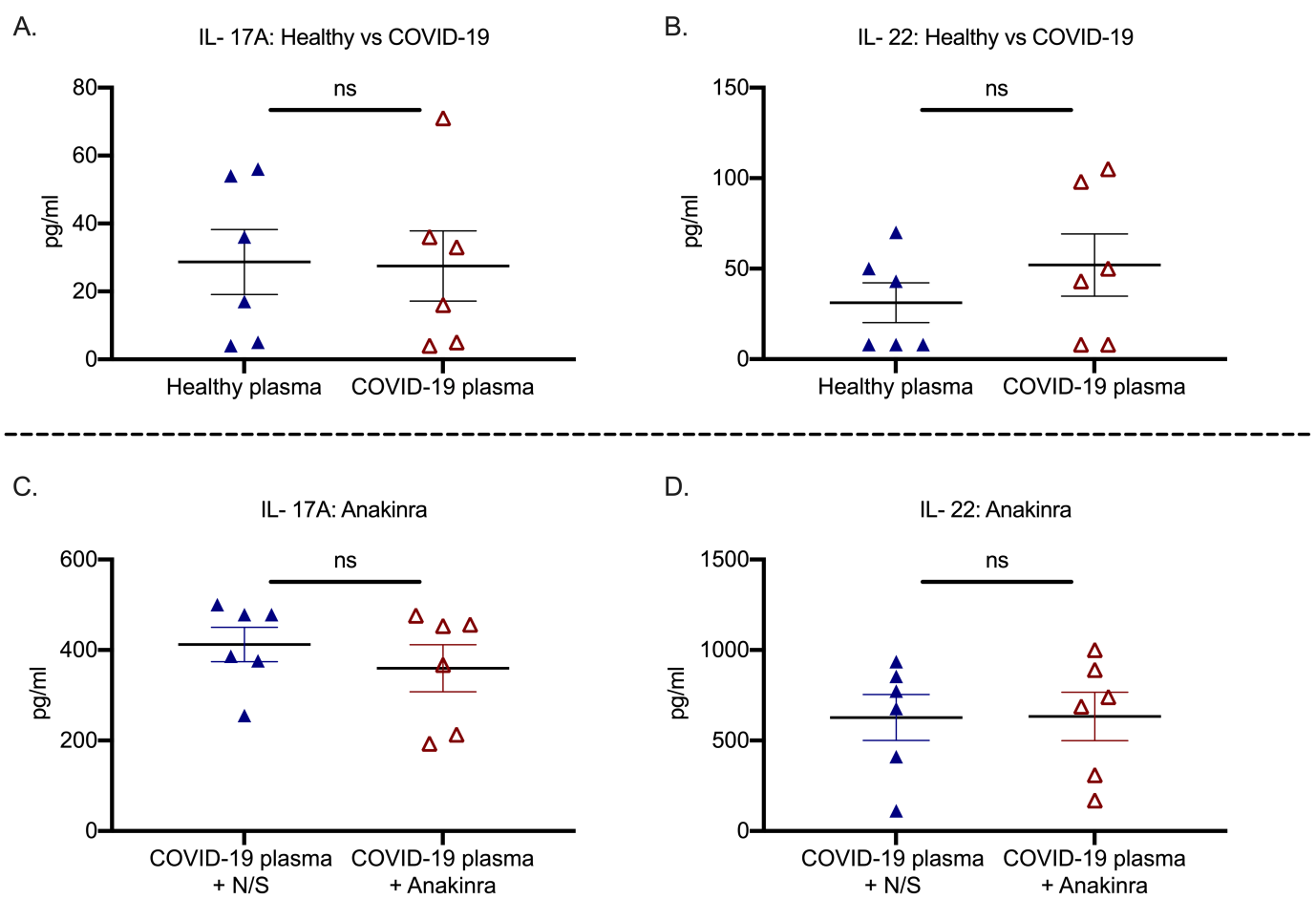

**Figure S1 IL-1 inhibition does not affect Th17 responses in COVID-19**

A-B) In a COVID- like infection model, C57Bl6 mice were treated intravenously (i.v.) with plasma of healthy volunteers or patients with ARDS due to COVID-19 for three consecutive days.

C-D) In a COVID- like infection model, C57Bl6 mice were treated intravenously (i.v.) with plasma of patients with ARDS due to COVID-19 with or without treatment with the IL-1 receptor inhibitor anakinra for three consecutive days.

Mice were sacrificed on day 4. Splenocytes were isolated and incubated with C. albicans. Th17 response is shown. Comparison by the Mann Whitney U test; ns non-significant. Treatment of mice with anakinra did not change the ex vivo production capacity of IL- 17A or IL- 22 indicating a lack of correlation with Th17 responses.

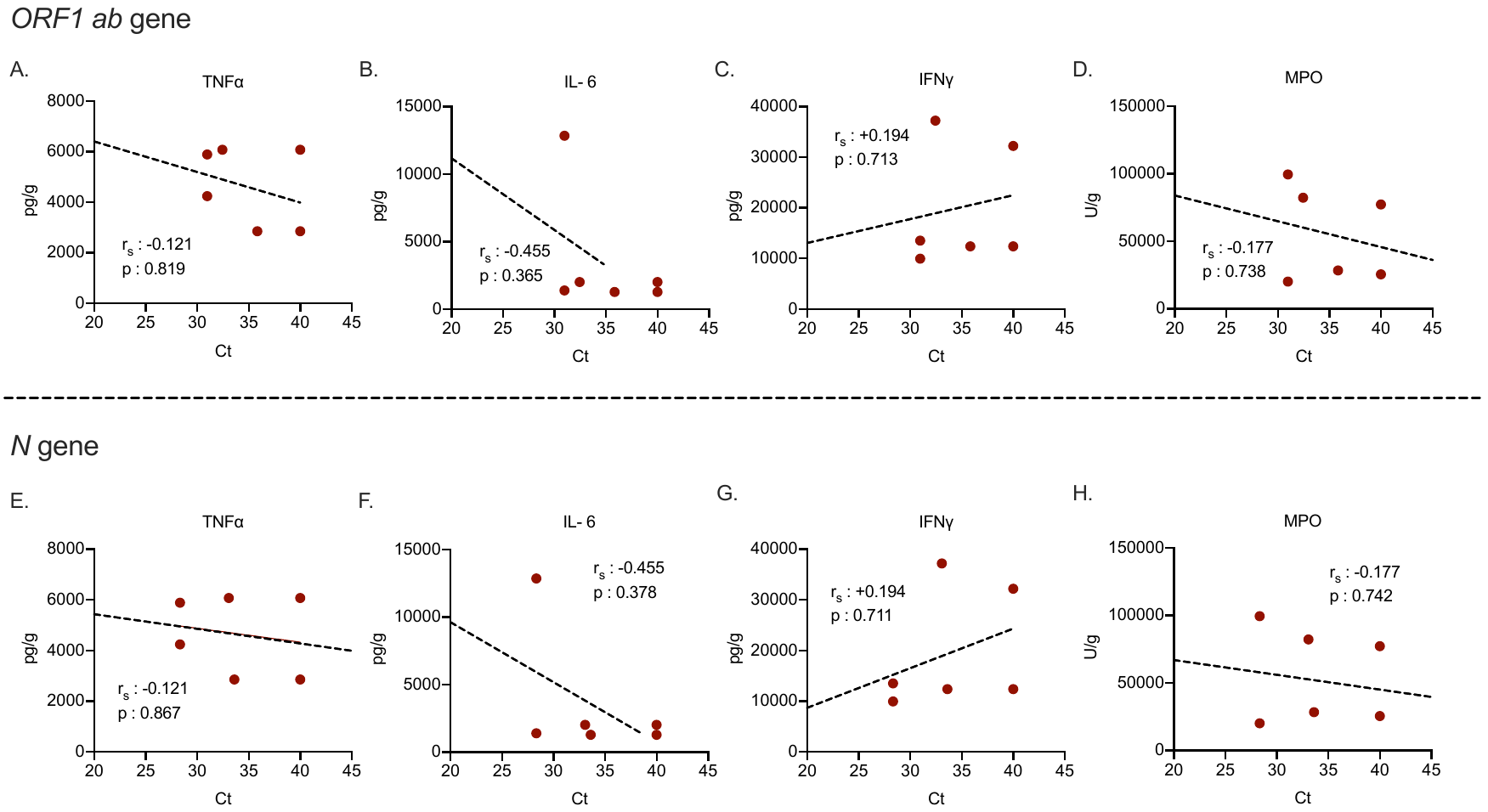

**Figure S2 Lack of correlation between cytokine production in tissues and number of viral transcripts**

The number of transcripts of the *ORF1 ab* gene and *N* gene of SARS-COV-2 was determined in plasma of patients with ARDS due to COVID-19. In a COVID- like infection model, C57Bl6 mice were treated intravenously (i.v.) with the same plasma from patients with ARDS due to COVID-19 for three consecutive days. Correlation of number of transcripts of the ORF1 ab gene (Α – D) and n gene (E – H) with TNFα, IL- 6, IFNγ and MPO in the lung. Spearman rank correlation coefficients (r_s_), interpolation lines and p- values are given.

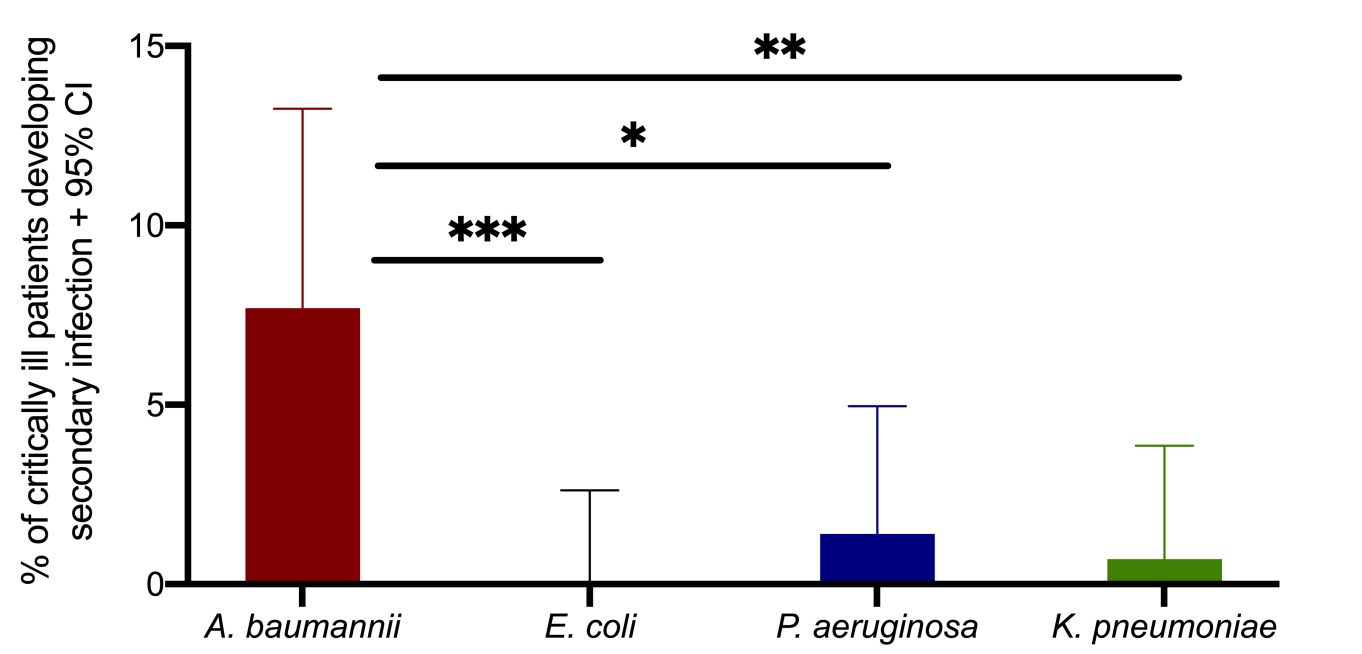

**Figure S3 COVID- 19 is associated with increased susceptibility to secondary infection due to *A. baumannii***

Percentage of critically ill COVID- 19 patients who developed secondary bacteremia by Gram- negative bacteria within 28 days after admission, sorted by type of bacteria. Comparisons by the Fisher exact test. ^✱^ p< 0.05; ^✱✱^ p< 0.01; ^✱✱✱✱^ p< 0.0001.

| **Table S1. Baseline clinical and laboratory characteristics of critically ill patients with pneumonia by SARS-CoV-2 coronavirus according to development of secondary bacteremia or not** | | | | |
| --- | --- | --- | --- | --- |
|  | *A. baumannii* bacteremia  (n=11) | Other gram-negative  bacteriemia  (n=3) | No secondary bacteremia  (n=120) | p-value |
| Age (years, mean ± SD) ^#^ | 66.11 ± 9.75 | 63.00 ± 1.73 | 63.92 ± 13.57 | 0.884 |
| CCI (mean ± SD) ^#^ | 3.00 ± 1.80 | 2.33 ± 0.58 | 2.79 ± 1.81 | 0.854 |
| APACHE II score (mean ± SD) ^#^ | 11.89 ± 5.01 | 9.33 ± 5.25 | 10.26 ± 5.18 | 0.663 |
| SOFA (mean ± SD) ^#^ | 5.67 ± 1.87 | 6.00 ± 2.65 | 6.10 ± 2.88 | 0.907 |
| Laboratory values (mean ± SD) | | | | |
| White blood cells (mm^3^) | 13620 ± 5085 | 11777 ± 3487 | 10095 ± 5252 | 0.206^#^ |
| Absolute neutrophil counts (mm^3^) | 11950 ± 4794 | 10695 ± 3678 | 8522 ± 4956 | 0.167^#^ |
| Absolute lymphocyte counts (mm^3^) | 951 ± 739 | 689 ± 219 | 913 ± 629 | 0.819^#^ |
| Platelets (x 10^3^, mm^3^) | 289428 ± 65739 | 263000 ± 32741 | 253731 ± 108144 | 0.683^#^ |
| C-reactive protein (mg/l) | 154.72 ± 123.90 | 142.77 ± 105.96 | 137.54 ± 215.62 | 0.984^#^ |
| Ferritin (ng/ml) | 1502 ± 1844 | 815 ± 566 | 1439 ± 1786 | 0.829^#^ |
| Procalcitonin (ng/ml) | 042 ± 0.65 | 0.13 ± 0.05 | 0.98 ± 1.96 | 0.694^#^ |
| D-dimers (ng/ml) | 18571 ± 38008 | 1046 ± 631 | 3715 ± 16917 | 0.222^#^ |
| AST (U/l) | 49.21 ± 63.40 | 48.00 ± 34.66 | 51.51 ± 41.31 | 0.982^#^ |
| ALT (U/l) | 35.80 ± 23.75 | 44.33 ± 24.38 | 52.68 ± 57.96 | 0.727^#^ |
| Creatinine (mg/dl) | 0.81 ± 0.33 | 0.77 ± 0.06 | 1.26 ± 1.60 | 0.667^#^ |
| Abbreviations: SD: standard deviation; APACHE: acute physiology and chronic health evaluation; CCI: Charlson’s comorbidity index; SOFA: sequential organ failure; AST: aspartate aminotransferase; ALT: alanine aminotransferase.  Comparisons by one-way ANOVA with the Bonferroni correction. | | | | |
